# Multicenter pivotal trial of the Minima stent for vascular stenosis in infants and young children

**DOI:** 10.1101/2025.06.19.25328682

**Authors:** Patrick M Sullivan, Evan M Zahn, Shyam Sathanandam, Brian Morray, Shabana Shahanavaz, Arash Salavitabar, Aimee K. Armstrong, Diego Porras, Darren P Berman

## Abstract

**Background:** The Minima Stent System is the first stent designed, tested, and FDA-approved for use in neonates, infants, and children. Our objective was to evaluate the safety and efficacy of Minima implantation for pulmonary artery stenosis (PAS) and coarctation of the aorta (CoA).

**Methods:** Multicenter, single-arm, prospective, non-randomized trial. Primary endpoints included stenosis relief, freedom from device-related serious adverse events (SAEs) or surgical intervention through 6-months, and maintenance of vessel lumen diameter on computed tomography or catheter angiography at 6-months.

**Results:** Forty-two patients (21 PAS, 15 recurrent CoA, 6 native CoA) underwent Minima implantation at a median age and weight of 9 (range 0.4-112) months and 7.8 (3.4-28.3) kg. Implantation was successful in 41 (97.6%) and resulted in a median increase in minimal vessel diameter of 131% (46-483%) and reduction in median pressure gradients from 25 (0-63) mmHg to 0 (0-6;p<0.001) mmHg in CoA patients. Two acute PAS stent embolizations occurred; both stents were secured in the contralateral PA, and one was treated with an additional Minima stent. Seven CoA patients, all under 6kg, experienced transient femoral artery thrombosis. At 6-months there were no explants or device-related SAEs. Luminal diameter was maintained at 89% (59-137%) of implant diameter. During a median follow-up of 596 (412-979) days, 13 (31%;7 CoA, 6 PAS) patients underwent planned stent expansion without complications.

**Conclusions:** The Minima system is safe and effective for treating PAS and CoA in infants and small pediatric patients. Luminal patency was preserved, and planned re-interventions for somatic growth seem well tolerated in early follow-up.

**What Is Known?:** Endovascular stent therapy for infants and small children with congenital heart disease has remained highly challenging due to a dearth of available tools specifically designed, tested, and approved for use in this unique population.

**What the Study Adds.:** The Minima Stent System, specifically designed for infants and small children, has demonstrated safety and effectiveness in treating both native and post-surgical coarctation of the aorta and pulmonary artery stenosis in this population. On follow-up imaging, vessel patency and caliber were well-preserved, and further elective expansion of the stent to accommodate somatic growth was well-tolerated. Additional follow-up of the current cohort and other post-approval implants is needed to better understand the long-term performance of the Minima stent as patients approach adult size.

## Introduction

Since first described nearly 35 years ago ^1, 2^, transcatheter stent implantation has become the preferred therapy for older children, adolescents, and adults with congenital heart disease (CHD) and pulmonary artery stenosis (PAS) ^3, 4^ or coarctation of the aorta (CoA) ^5, 6^. Although several stents are approved for use in larger pediatric and adult patients with CHD, ongoing challenges have limited the development of stents designed specifically for treating infants and small children with PAS and CoA ^7, 8^. This has led to widespread “off-label” use of stents intended for adults with acquired cardiovascular disease in this unique and challenging population ^9–13^. In addition to being untested in the pediatric population, these devices tend to lack features ideal for use in small patients.

The Minima Stent System™ (Renata Medical, Newport Beach, CA) was developed to meet the unmet need for a low-profile transcatheter pediatric stent with the capacity to reach adult vessel size through serial expansion. A multicenter early feasibility study (EFS) assessing the performance, safety, and efficacy of Minima stent implantations in infants and small children with PAS and CoA demonstrated promising results ^14^. Based upon the EFS results, enrollment was expanded to a larger pivotal trial, the results of which are reported herein.

## Methods

### Study design

This multicenter, single-arm, prospective, non-randomized trial (ClinicalTrials.gov ID:NCT05086016) was open to 42 neonates, infants, and children with native or recurrent CoA or branch PAS. Seven centers in the United States were included, and the study was reviewed and approved by each institutional review board. Subject inclusion/exclusion criteria and safety and efficacy endpoints are listed in **Supplemental Table 1**. A patient was considered enrolled in the study after informed consent was signed for participation and the procedure to insert the stent had been initiated, defined as the time of vascular access.

The schedule for assessments and follow-up testing is outlined in our previous publication of the EFS results ^14^. After the EFS was completed, the follow-up schedule was modified to include a cardiac computed tomography angiogram (CTA) or catheter angiography at six months post-implant to evaluate the primary outcome measure of maintenance of stent lumen caliber. Sites reported inner lumen minimal diameter measurements of the stented vessels. A stented vessel with a minimal lumen diameter <50% of the post-implant lumen diameter on 6-month follow-up angiography and no other obvious mechanism of stent dysfunction, like fracture or deformation, was considered to have significant in-stent restenosis. The ten subjects enrolled in the EFS were re-consented and were included in the pivotal cohort. An independent Data Safety Monitoring Board reviewed procedural results and adverse events on a quarterly basis.

### Renata Minima stent system and procedure details

The Minima stent is a 17 mm length closed-cell, balloon-expandable, cobalt-chromium stent designed to maintain structural integrity and radial strength over a range of achievable diameters from 5.1 mm to 24 mm (**Figure 1**). Using standard balloon angioplasty tools and techniques, the stent may be serially expanded to keep pace with somatic growth and has a predictable amount of foreshortening as it is expanded ^15^. Currently, the stent comes pre-mounted on either a 6 mm or 8 mm semi-compliant balloon allowing for initial implant diameters between 5.1 mm and 8.5 mm, depending on the pressure delivered to the balloon. The kink-resistant delivery system has a 63 cm working length, the same outer diameter as a 4 French sheath, and a retractable cover over the stent that ensures a smooth transition to the balloon tip, which serves as a tapered dilator that can be directly introduced through the skin or through a 6 Fr sheath over a 0.014” or 0.018” wire.

**Figure 1:**
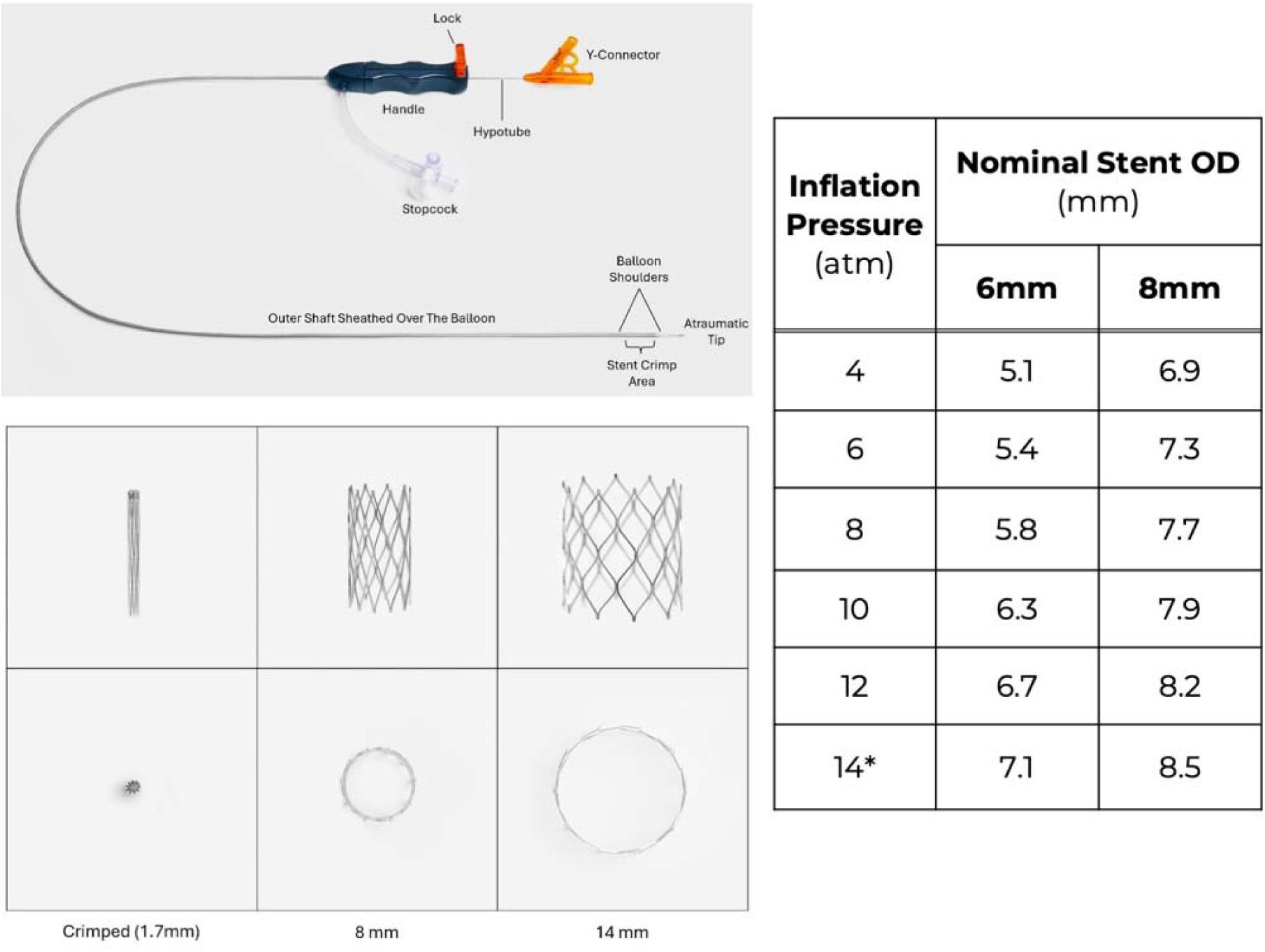
Renata pre-mounted Minima delivery system (above) with crimped and serially expanded stent (below) and range of delivery diameters (right).

Procedural details for initial implantation have been described previously ^14^. Angiographic measurements made during the implant procedure were confirmed by a core lab that did not otherwise participate in the study. Patients were considered for re-dilation of the Minima stent at the discretion of the implanters. Re-dilation procedures were performed using standard angioplasty tools and methods to serially expand the stent in 2 mm increments, as required in the protocol. Re-dilations were deemed effective if the achieved stent luminal diameter was within 2 mm of the adjacent vessel diameter.

### Data analysis

Baseline demographic, clinical, procedural, follow-up, imaging, and re-dilation data were collected prospectively. Descriptive continuous variables are presented as median (range) and categorical variables as n (%). Comparisons of continuous variable characteristics were tested using 2-sample t-tests with unequal variance. Follow-up data are reported through October 7, 2024. The Kaplan-Meier method was employed to estimate freedom from re-intervention for the cohort and among subgroups. Univariate Cox proportional hazards regression models were used to assess for associations between freedom from re-intervention and selected variables. P-values <0.05 were considered statistically significant.

## Results

### Study cohort

Forty-two patients underwent attempted Minima stent implantation between February 2022 and August 2023 at an age and weight of 9 (0.4-112) months and 7.8 (3.4-28.3) kg, respectively (**Figure 2**). The 6 mm balloon system was used in 16 patients (38%), and the 8 mm system was used in 26 patients (62%). Eleven additional patients were consented for enrollment and underwent catheterization, but the Minima stent was not opened due to findings at the time of catheterization, including: vessel diameter or length outside of sizing criteria (n=8), successful balloon angioplasty (n=1), hemodynamics that did not warrant intervention (n=1), and stenosis not located in study target location (n=1).

**Figure 2:**
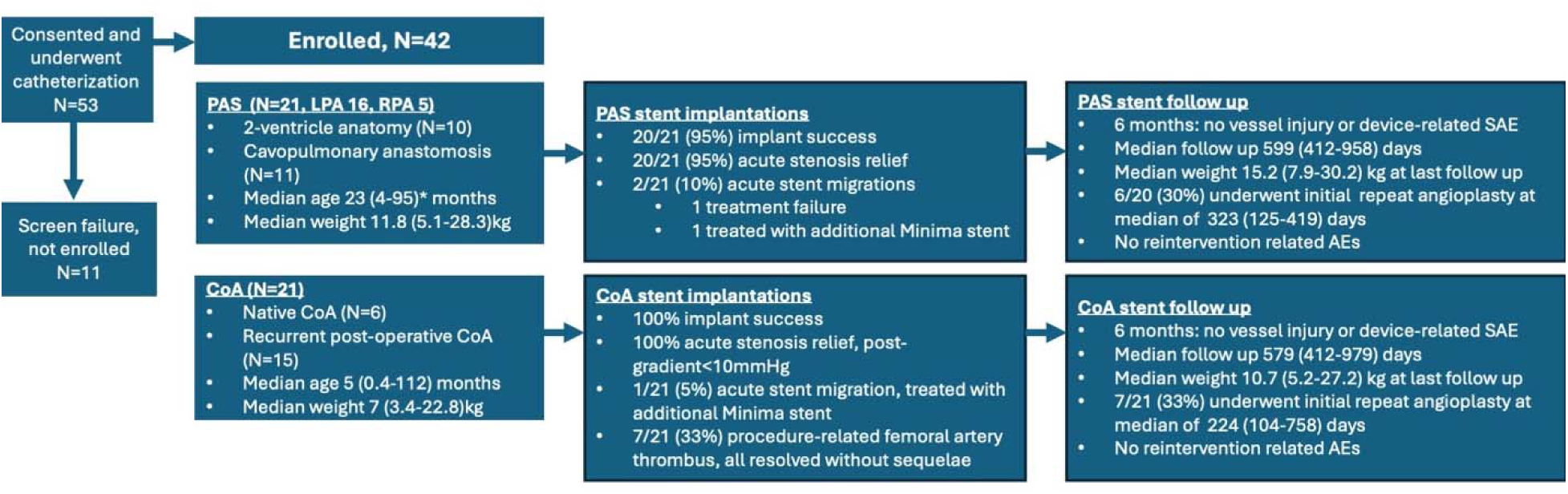
Flowchart of enrollment and follow up. Abbreviations: PAS (pulmonary artery stenosis), CoA (coarctation of the aorta), AE (adverse event). *All data presented as median (range)

Baseline demographic and clinical data for the cohort are provided in **Table 1**. The majority were under 24 months of age (71.4%) and weighed under 10 kg (64.3%). Twenty-one patients (50%) were enrolled for treatment of CoA at an age and weight of 5 (range 0.4-112) months and 7.0 (range 3.4-22.8) kg, including 6 with native CoA. The remaining 21 patients (50%) were enrolled for treatment of PAS at an age and weight of 23 (range 4-95) months and 11.8 (range 5.1-28.3) kg.

**Table 1:**
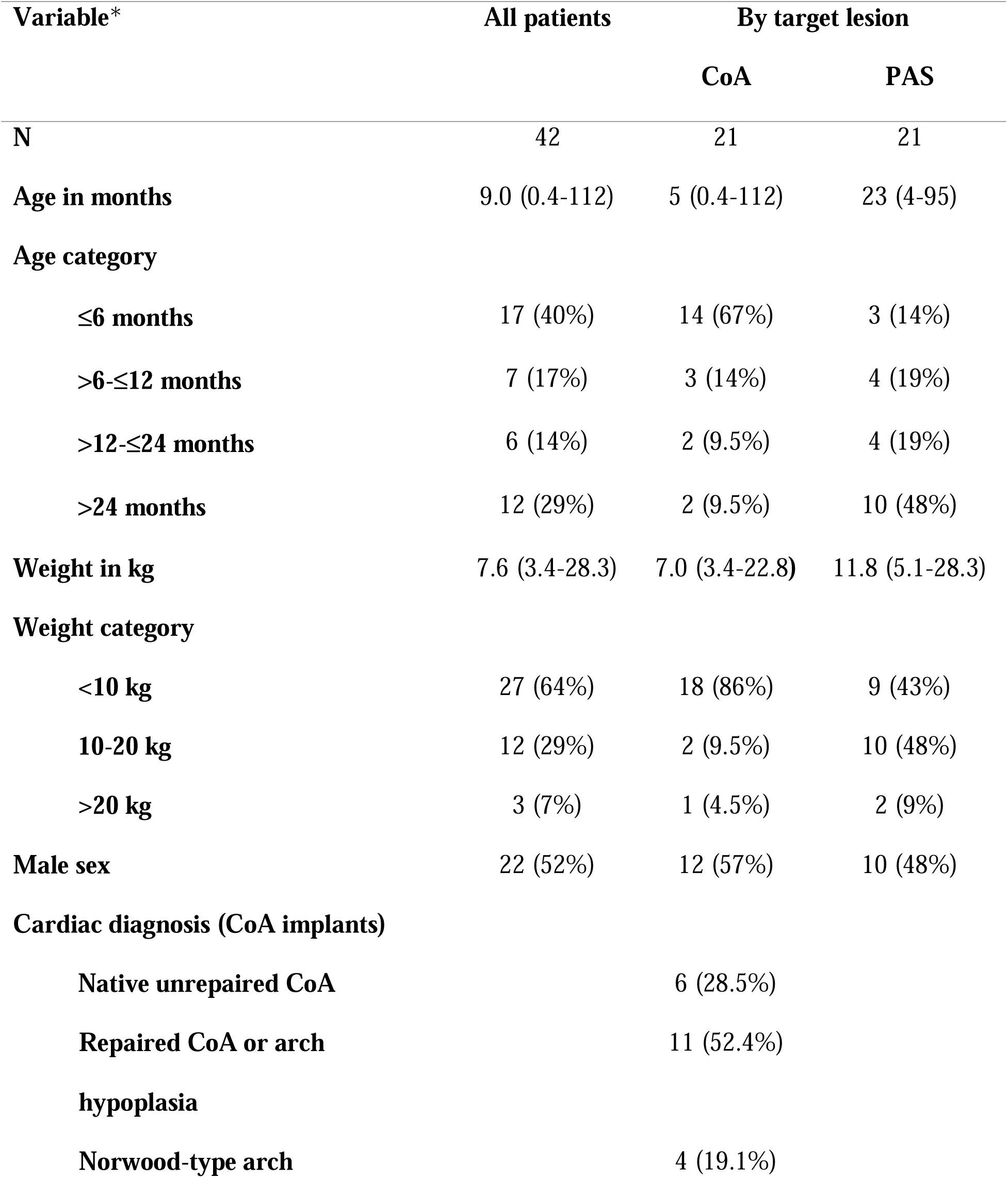
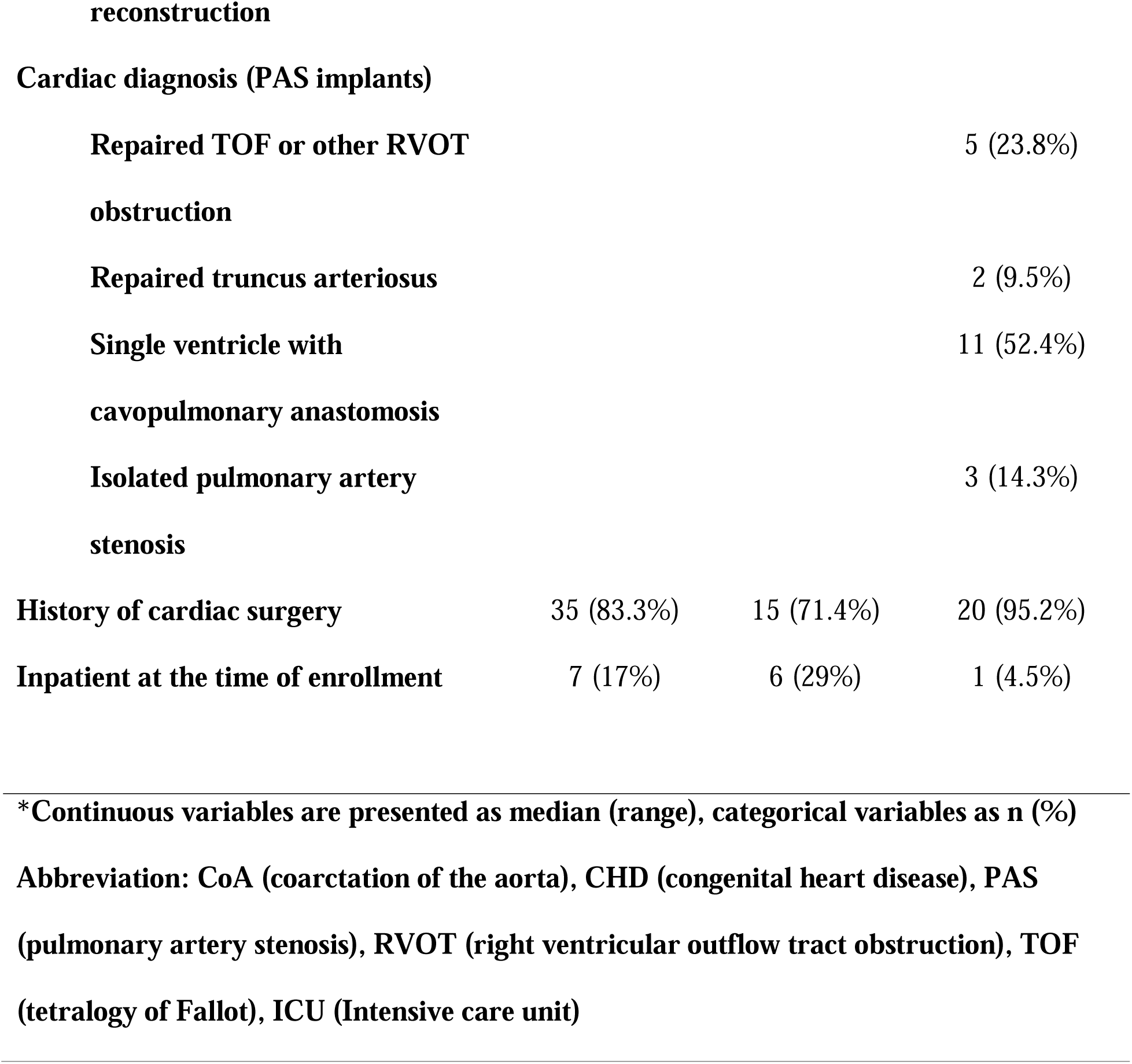
Cohort demographic and clinical characteristics

### Implant procedure

Successful implantation of the Minima stent in the intended location occurred in 41 of the 42 patients (97.6%). Thirty-nine patients (92.8%) underwent implantation of one Minima stent, and three patients underwent implantation of two Minima stents, resulting in a total of 45 implantations. Acute stenosis relief was achieved in all 41 patients who successfully underwent implantation. The minimal vessel diameter increased by 131% (46-483%), by 3.6 mm (1.5-6.6 mm), and reached 98% (71-162%) of the surrounding vessel size.

Among CoA stent implants, invasive systolic pressure gradients improved from 25 mmHg (0-63 mmHg) to 0 mmHg (0-6 mmHg; p <0.001; **Figure 3A**). Among the PAS stent implants in patients with biventricular circulation and available pressure data (N=9 for pre-implant gradient, N=5 for both pre- and post-implant gradient), systolic pressure gradients improved from 29 mmHg (2-50 mmHg) to 5 mmHg (0-10 mmHg; p=0.007).

Two PAS patients with left pulmonary artery (LPA) ostial stenosis experienced acute stent dislodgment into the main pulmonary artery during withdrawal of the delivery balloon after deployment. In one case, the stent had been deployed more proximally than intended, whereas in the other, the distal vessel diameter fell outside the sizing criteria necessary to ensure adequate stent-vessel wall apposition. Both stents were redirected and redeployed within the contralateral pulmonary artery. In the first case, an additional Minima stent was successfully deployed in the target lesion. In the other case, which marked the only treatment failure in the cohort, further stent treatment of the lesion was deferred.

In two additional PAS patients, the unsheathed stent was noted to move on the delivery balloon prior to inflation. In both cases, the cause was attributed to mechanical interaction with adjacent stents placed during a previous procedure. In one case, the Minima stent was successfully deployed distal to the target lesion, and an additional Minima was placed successfully across in the lesion. In the other case, the Minima stent was successfully deployed in the lesion. One patient with CoA required an additional Minima stent after the initial stent shifted slightly proximal during angiographic catheter manipulation following deployment but remained within the lesion. There were no post-procedure stent migrations.

Seven CoA patients (33%) with a median weight of 5.1 kg (3.6-5.9 kg) had ultrasound evidence of procedure-related femoral artery thrombosis. Six were treated with enoxaparin, and all resolved without ongoing sequelae. One of these patients developed a pseudoaneurysm and a small arteriovenous fistula at the access site, which resolved on follow-up imaging. No additional significant procedural adverse events were reported.

### Follow-up and re-interventions

At six months of follow-up, there were no deaths or open surgical interventions due to stent dysfunction. Additionally, there were no reported device- or procedure-related SAEs and no evidence of stent fracture or late migration. No tricuspid valve injuries were noted in biventricular PAS patients on follow-up. Among the 41 patients who underwent successful stent implantation, 37 had follow-up CTA six months after implantation, while four had catheter angiography between 104 and 281 days after initial implantation instead of CTA (**Figure 3-5**).

**Figure 3:**
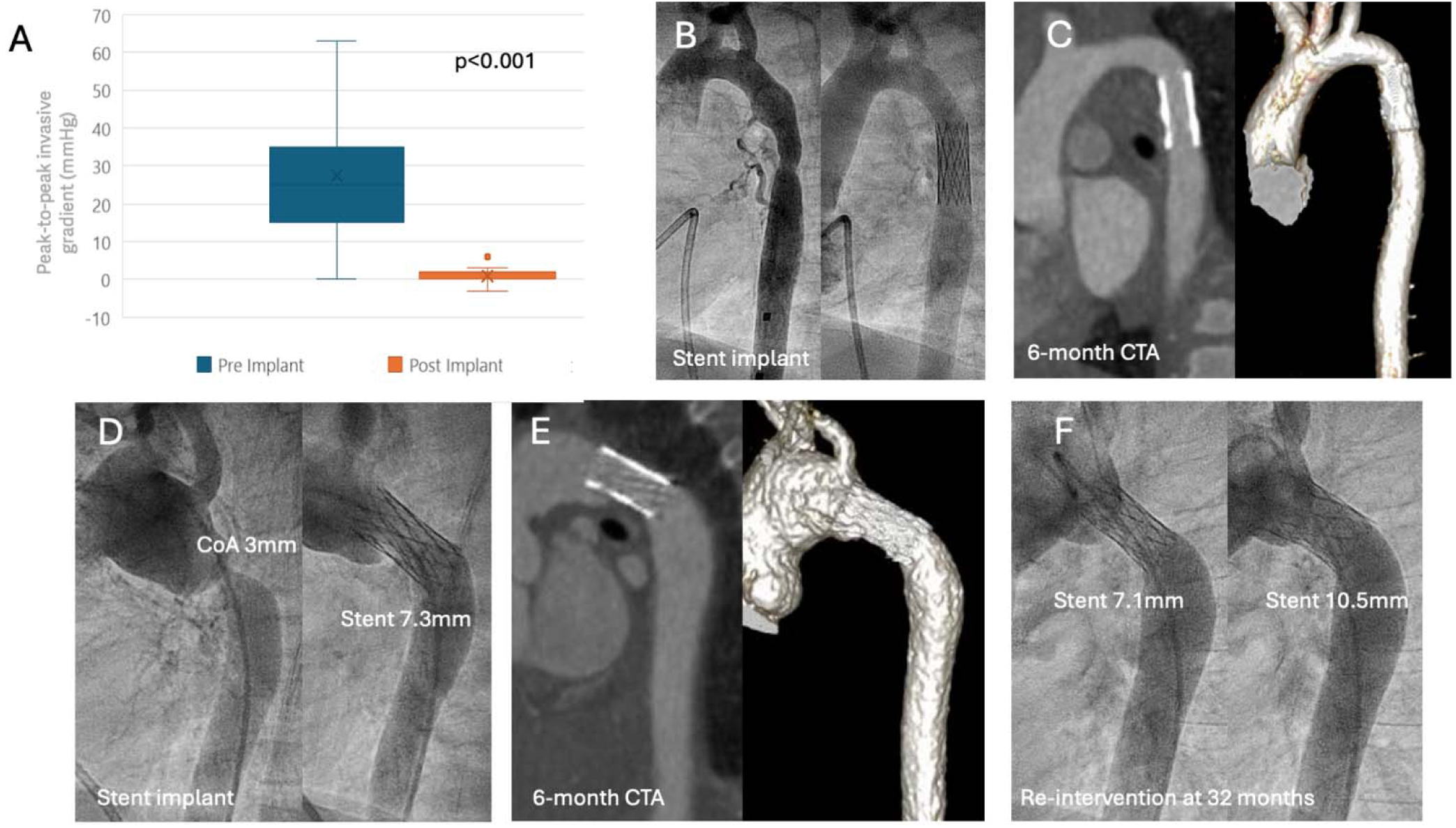
Minima therapy for CoA. (A) Stenting resulted in reduced aortic gradients. (B) Pre- (left) and post-intervention (right) angiograms show stenosis relief in an 8.4 kg infant with native CoA and (C) CTA images 6 months later show maintained stent patency and no aortic wall injury. (D) Similar pre- and post-implantation angiograms are shown for a 5.5 kg infant with recurrent CoA with (E) 6-month follow-up CTA. (F) At 32 months after implant, stent caliber is stable (left) and improved after re-intervention (right) with serially larger balloons.

**Figure 4:**
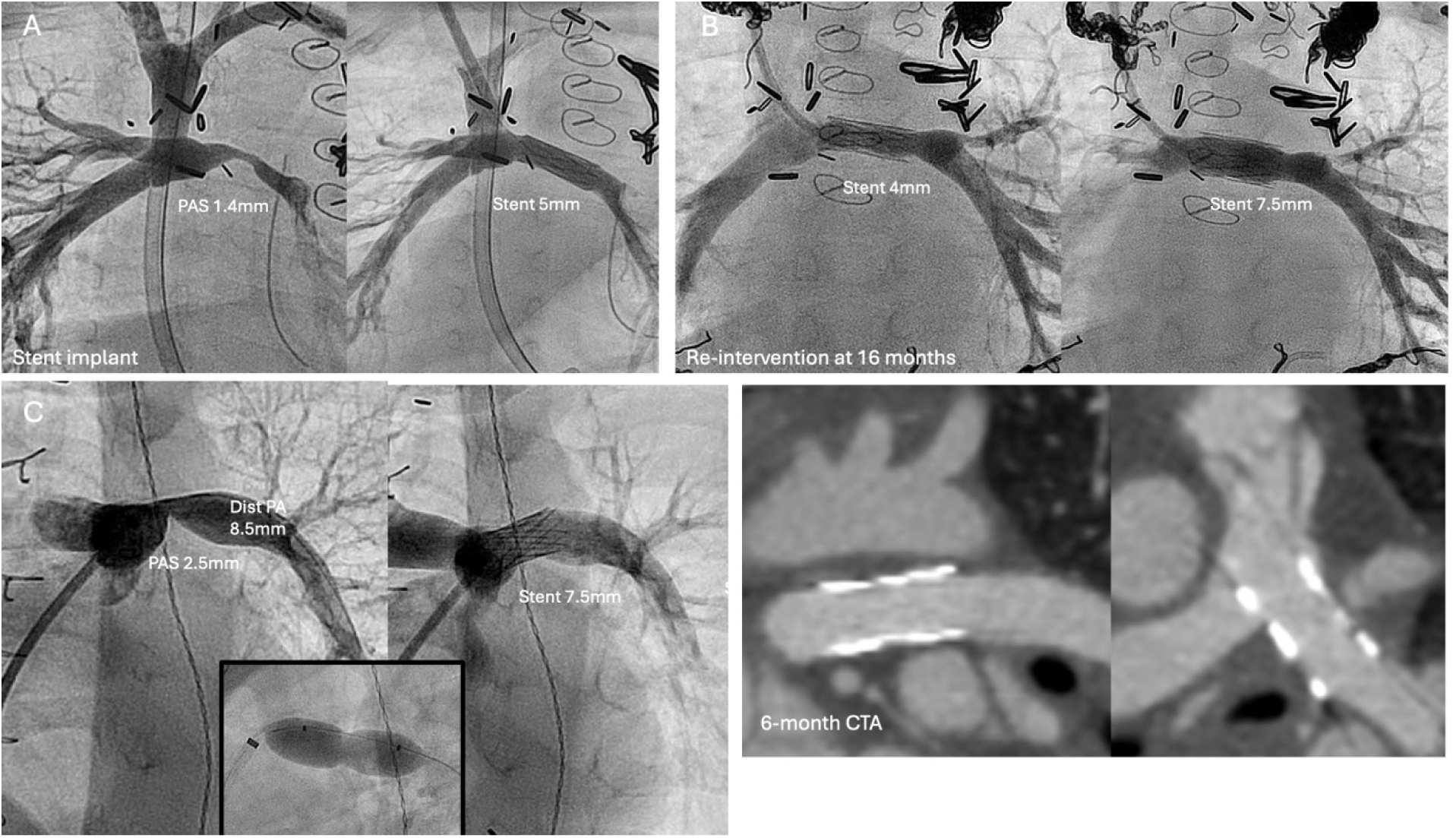
Minima therapy for PAS. (A) Pre- (left) and post-implantation (right) angiograms in a 6 kg infant with superior cavopulmonary shunt show relief of severe left pulmonary artery (LPA) stenosis. (B) At re-dilation for somatic growth 16 months later, implant caliber is stable (left) and further dilation (right) keeps pace with distal vessel growth. (C) Similar pre-(left) and post-implantation (right) angiography in a 14kg male with repaired tetralogy of Fallot and LPA ostial stenosis after balloon compliance testing (middle inset). (D) CTA at 6-months shows maintained patency and caliber of the stent.

**Figure 5:**
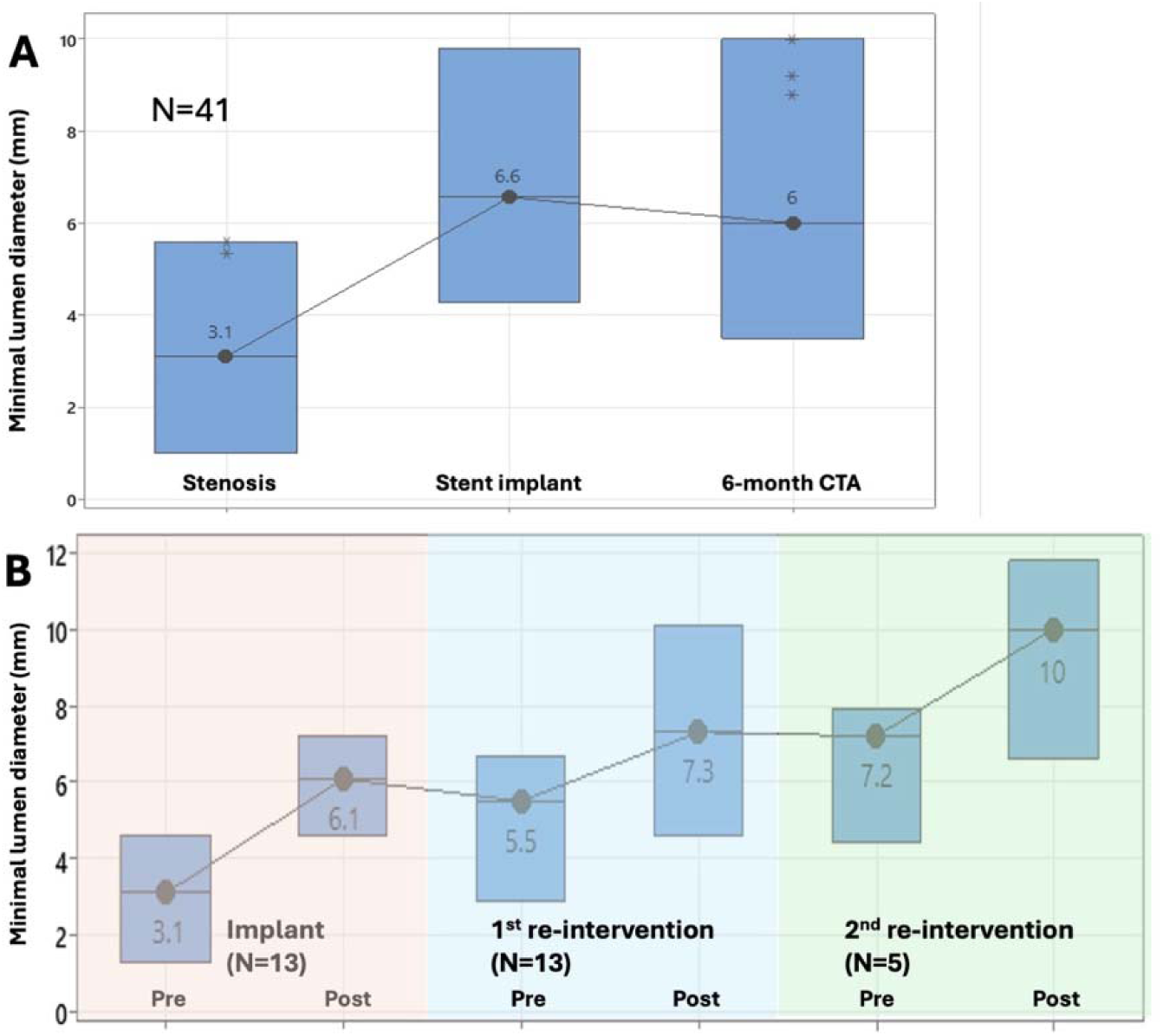
Stent lumen diameter at initial implantation and follow-up. (A) Minima implants relieved stenosis and maintained vessel caliber at 6-months follow-up. (B) Among the 13 subjects who underwent at least one re-dilation and five who underwent additional re-dilation, vessel diameters reflect growth with each dilation and maintained caliber from one intervention to the next. Box plots depict median and range of values.

Intraluminal vessel diameters were maintained at a median of 89% (59-137%) of the initial implant diameter, and no stent deformation, stent fractures, or stent-related vessel wall injuries were identified. Notably, two patients underwent elective reintervention prior to 6-months of follow-up. Both underwent catheterization for indications other than Minima re-dilation but were noted during catheterization to have somatic growth that would accommodate further stent dilation. One patient with a Minima stent in the aortic position underwent stent explant at 224 days post-implant during a planned cardiac operation to address several other surgical lesions, as future stent dilations were felt to place the left bronchus at risk for compression.

During a median follow-up period of 596 days (412-979 days), the weight of the cohort increased from 7.8 kg (3.4-28.3 kg) to 13.4 kg (5.2-30.2 kg). Thirteen patients, seven with CoA and six with PAS, underwent at least one additional stent dilation at a median of 281 days (104-758) from implant. Among these patients, the weight at re-intervention was 10.4 kg (6.8-15.2 kg) compared to 6.2 kg (3.9-11.9 kg) at initial stent implantation. The freedom from reintervention was 95% [95%CI: 82-99%], 81% [95%CI: 66-90%], and 70% [95%CI: 53-82%] at 6 months, 1 year, and 2 years of follow-up, respectively. The stated indication for re-intervention was the growth of the surrounding vessel noted on follow-up imaging or catheterization in 11 cases and mean echo Doppler gradient ≥20mmHg across the stented vessel in the remaining 2 cases. The vessel lumen diameters prior to further balloon expansion at the time of re-intervention measured a median of 84% (56-100%) of the initial post-implant lumen diameter with no lumens measuring <50% of implant caliber. No procedure-related adverse events were reported during these re-interventions.

The weight at the time of initial stent implantation was lower in patients who underwent re-dilation within one year of follow-up (5.9 kg (3.9-10.7 kg), p=0.001) or at any point during follow-up (6.2 kg (3.9-11.9 kg), p=0.006) compared to those who did not undergo re-dilation procedures (8.7kg (3.4-28.3)). Weight less than the cohort median approached but did not meet statistical significance for increased risk of earlier re-intervention (HR 2.8 [95%CI 0.9-9.3], p=0.08; **Figure 6B**). However, higher weight using weight at initial implant as a continuous variable was associated with longer time to re-intervention (HR 0.8 for each 1 kg increase in weight [95%CI 0.7-0.99], p=0.04; **Table 2**). The time to re-intervention trended towards being longer among those who underwent initial stent implantation with the 8mm system compared to the 6mm system (p=0.09; **Table 2**, **Figure 6B**). Time to re-intervention was similar between CoA and PAS stent implants (p=0.9, **Table 2**, **Figure 6C**).

**Figure 6:**
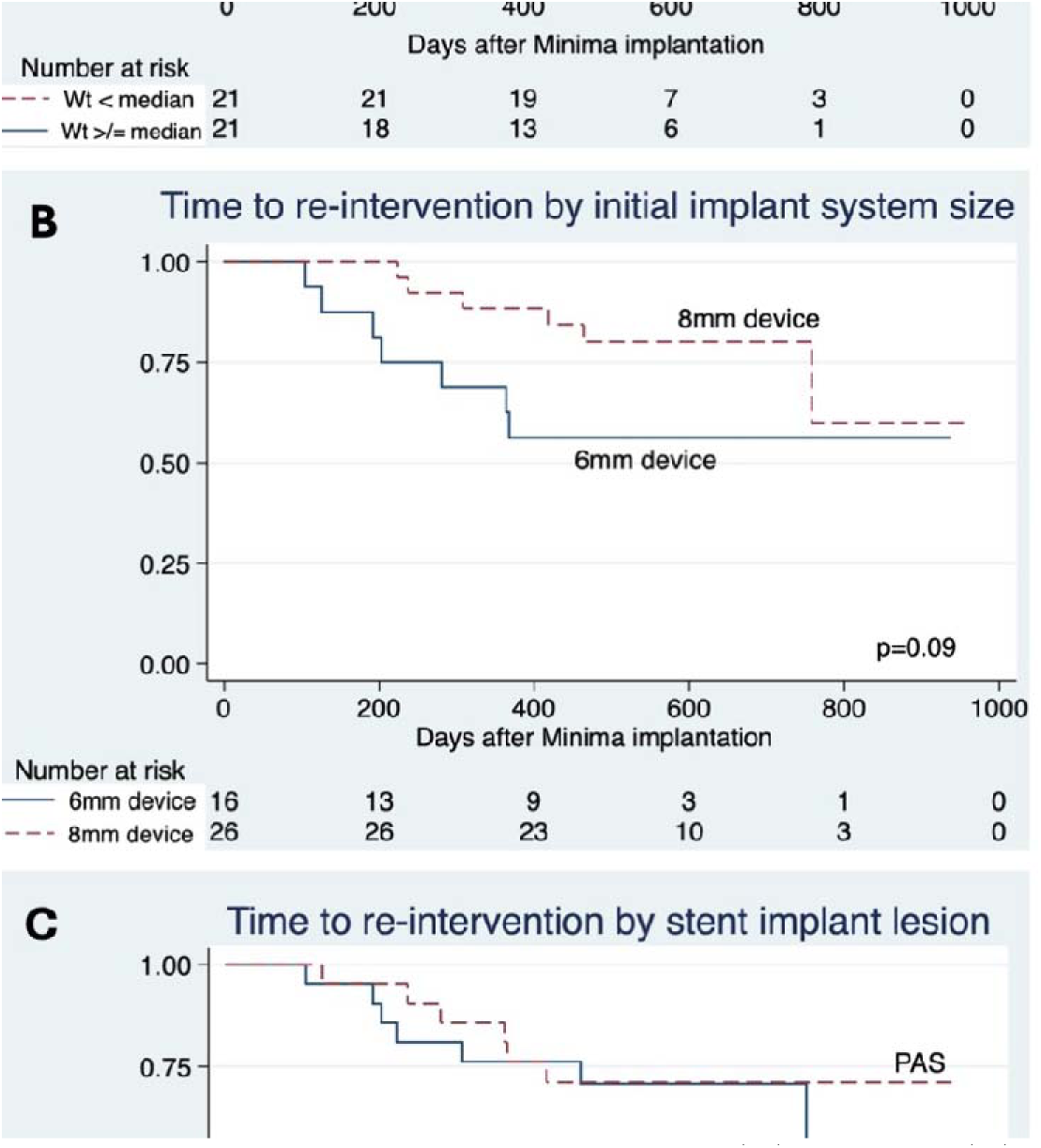
Freedom from transcatheter re-intervention stratified by (A) weight, (B) initial implant system, and (C) lesion type. P-values were derived from univariate Cox proportional hazards regression.

**Table 2:**
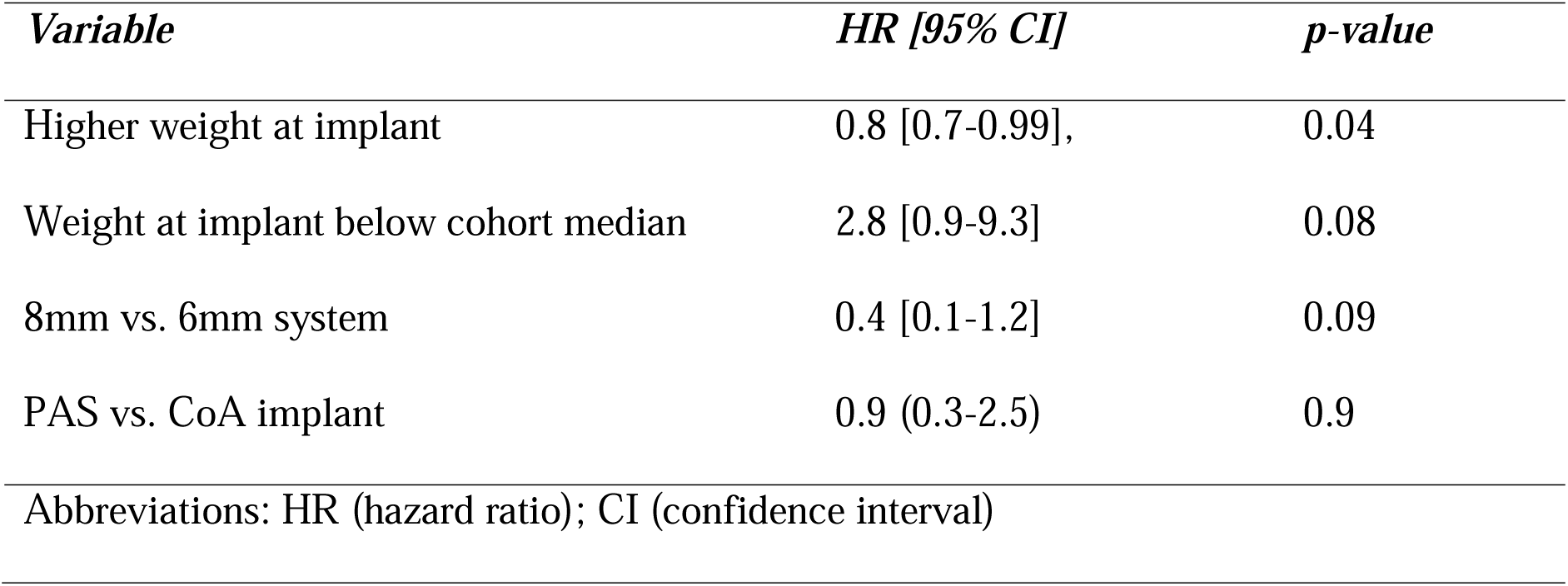
Proportional hazards regression of risk factors for earlier re-dilation

Initial re-dilations (N=13) utilized median maximal balloon sizes that were 2 (0-4) mm larger than the initial implant balloon, resulting in an increase in the minimal stent lumen diameter from 5.5 mm (2.9-6.7 mm) to 7.3 mm (4.6-10.1 mm; p=0.002). Second re-dilations (N=5) utilized maximal balloon sizes that were 4 (2-4) mm larger than the initial implant, further increasing the minimal stent lumen diameter from 7.2 mm (4.4-7.9 mm) to 10 mm (6.6-11.8 mm; p=0.06, **Figure 5B**). All stents undergoing re-dilation were expanded to within 2 mm of the surrounding vessel size except for two cases in which the operators chose to undersize the balloon relative to the neighboring vessel due to unique anatomical considerations. No additional stents were implanted during re-interventions.

## Discussion

Herein, we report the results of a multicenter pivotal trial using the novel Minima stent system to treat CoA and PAS in infants and small children. In a heterogenous cohort of 42 patients, half with either native or post-operative recurrent CoA and half with PAS, the Minima stent was successfully implanted in all but one patient achieving safe and durable relief of vascular obstruction. Follow-up multiplanar imaging demonstrated well-maintained lumen calibers and stent integrity without vessel wall injuries. Based on these findings, in August 2024 the Minima stent became the first stent approved by the FDA for use in small pediatric patients. During follow-up ranging from 13 to 32 months, 31% of the cohort underwent elective re-dilation of the stent without any procedural adverse events.

The introduction of transcatheter stent therapy for vascular stenosis in the late 1980s was an important innovation for CHD patients ^1, 16^. In the ensuing decades, stent therapies have shown favorable safety and efficacy compared to balloon angioplasty and surgical intervention in older pediatric patients with pulmonary artery stenosis ^3, 17^, coarctation of the aorta ^5, 6^, and other lesions ^2, 18^. In pediatric patients who ultimately do require cardiac surgery, stent implantation has been shown to delay or reduce surgical interventions ^19, 20^. Despite the potential benefits of bespoke stent technologies designed specifically for CHD patients, challenges in device development, regulatory pathways, and marketplace realities, among other factors, have contributed to a general lack of available transcatheter stent technology to treat this unique population. ^7, 8^.

The lack of available pediatric stent therapies is most apparent in infants and small children, who pose unique anatomical challenges. While stent therapy has been performed in these small patients for years ^9–11^, this experience has relied on the creative off-label use of devices that are neither designed for, tested in, nor approved for use in this population ^12, 13^. This limitation has presented significant procedural hurdles and has restricted the number of small patients who benefit from stent implantation ^13^. Even when successfully deployed, off-label devices often lack essential features for use in smaller patients, such as low-profile delivery systems and the capacity to reach adult diameters through serial expansion to keep pace with somatic growth. Creative “workarounds” have been described, including purposeful fracturing ^21, 22^ of smaller stents as a child grows or modifying available larger stents to fit the anatomy of small patients ^23^; however, these approaches have drawbacks and have not become standard of care. Aside from having an appropriately small delivery profile and the capacity to reach adult vessel size, important agreed-upon features of the ideal pediatric stent ^24^ include trackability in small anatomy and through the heart, flexible and atraumatic design, and a structure that maintains radial strength and minimizes foreshortening with serial expansion over time.

The Minima delivery system profile was favorable for use in small vessels in the current cohort. Indeed, we report efficacious CoA stenting in patients as small as 3.4 kg. Vascular complications occurred expectedly at arterial access sites in patients weighing less than 6 kg but all resolved without longer term sequelae. Rates of femoral arterial injury in this group compare favorably with what has been reported in previous studies describing off-label aortic stenting and angioplasty in infants and other small patients ^25, 11^. While future design and procedural improvements may lower the risk of this complication, it is unlikely to be eliminated completely. Careful ultrasound-guided access techniques and implementation of arterial injury evaluation and treatment protocols will be necessary for minimizing the risk of CoA stent therapy in small patients ^26, 27^. The Minima delivery system was highly trackable over .014” and .018” wires, and delivery and deployment were well-tolerated. Particularly in PAS interventions in small patients, larger and stiffer systems designed for adult-sized stents can be difficult to deliver through the right heart and its valves and may cause hemodynamic instability or tricuspid valve injury during the intervention, none of which was observed in this cohort.

While this study demonstrated a high implant success rate with rare adverse events, the closed-cell cobalt-chromium Minima stent was notably more rigid than an open-cell stent design and less conformable to tightly angled vessels upon deployment. Although no patients were disqualified at the time of catheterization due to anatomical challenges, we speculate that the two LPA stent embolizations observed may have resulted in part from a combination of stent rigidity, relative under-sizing of the stent, and the acute angulation of the LPA take-off in these types of ostial and compliant stenoses. Treatment of ostial pulmonary artery stenosis presents unique challenges and carries a higher risk for stent embolization. In one pediatric series, 17% of ostial stents dislodged, and all but one required additional stent implantations for adequate lesion coverage ^28^.

While the Minima stent was designed and tested both in vitro and in a growing animal model to achieve adult vessel diameter ^15^, questions surrounding the long-term performance of the stent remain unanswered in this early study. Long-term data on off-label vascular stent implantations in small patients are limited to small single-center cohorts, and it is not clear what expectations we should have about the timing and frequency of re-interventions. For example, in a cohort of 27 patients undergoing implantation of various types and sizes of stents for PAS and CoA at a median age of 10 months, median time to re-intervention was 13 months, and all patients had required re-intervention by six years of follow-up ^10^. A more recent study of intraoperative stent implantations for PAS reported that 50% of patients implanted at less than two years of age required re-intervention by three years of follow-up ^17^. In a cohort of patients undergoing adult-sized stent implantations for CoA weighing under 20kg, 72% required re-intervention at a median of 49 months and freedom from re-intervention was 50% at two years of follow-up among patients weighing <10kg ^11^.

While these studies report the use of a variety of stent sizes, types, and delivery techniques, the rates and timing of re-intervention, particularly among the smallest patients, are comparable to the current cohort in whom the estimated freedom from re-intervention was 70% at two years of follow-up. Like the current study, these studies found that smaller initial patient size was associated with shorter time to re-intervention. This likely reflects the more rapid somatic growth experienced in infancy compared to later in childhood and the potentially greater impact of intimal ingrowth on stents implanted at smaller initial lumen diameters. In the current cohort, re-intervention with balloon angioplasty showed the expected stepwise increase in luminal diameter with each intervention and maintained luminal diameter in between interventions, consistent with long-term maintenance of stent integrity, limited in-stent restenosis, and progression towards adult vessel size.

## Limitations

While the evaluation of the Minima stent lumen at six months of follow-up was encouraging and suggested that stent patency was maintained during this time interval, the measurements relied on non-sedated CTA, which in some cases was limited by stent beam-hardening artifacts and may have resulted in suboptimal comparison measurements. Notably, in patients undergoing re-intervention, median catheter-derived lumen measurements prior to the further stent expansion were similar to CTA measurements in the cohort as a whole. The expectations for long-term outcomes, re-intervention, and expansion to adult vessel size are still poorly understood. Still, they will become clearer with longer follow-up time and the enrollment of more patients in a planned post-approval registry study.

## Conclusions

The Minima Stent System has demonstrated safety and efficacy in treating native and post-operative PAS and CoA in this cohort of infants and small children with CHD. On early follow-up, the stent maintained structural integrity and luminal patency, and planned further expansion for somatic growth was well-tolerated. Extra caution should be exercised in treating challenging and compliant lesions like LPA ostial stenoses, however. While there remains a substantial unmet need for technology designed for and tested in pediatric patients with CHD, the device is a promising addition for minimally invasive therapy in this unique population.

## Author disclosures

Patrick Sullivan: proctor/consultant for Renata Medical.

Evan Zahn: Medical director for Renata Medical; proctor/consultant for Abbott, B Braun, Edwards Lifesciences, Medtronic, WL Gore.

Shyam Sathanandam: Proctor/consultant, speaker’s bureau, research support for Abbott.

Brian Morray: proctor/consultant for Renata Medical, Abbott, Medtronic.

Shabana Shahanavaz: Proctor/consultant for Abbot, Edwards Lifesciences, WL Gore, Medtronic, Renata Medical

Arash Salavitabar: proctor/consultant for and research support from Renata Medical; research support from Medtronic and Edwards Lifesciences

Aimee Armstrong: proctor/consultant for Renata Medical, B. Braun, Edwards Lifesciences, Medtronic, Abbott; Research grant support from Edwards Lifesciences, Renata Medical, atHeart Medical, Starlight Cardiovascular; consultant for Occlutech and Cook Medical.

Diego Porras: proctor/consultant for Edwards Lifesciences

Darren Berman: proctor/consultant for Renata Medical, Abbott, B. Braun, Edwards Lifesciences, Medtronic.

No authors served as consultants or proctors for Renata Medical until after completion of this study and FDA approval of the Minima Stent System

## Data Availability

Data used to generate this manuscript will be available upon request.

## Acknowledgements

Dr. Sathanandam was affiliated with the Division of Pediatric Cardiology at the University of Tennessee Health Science Center in Memphis, TN during this trial and we would like to acknowledge this team for their efforts.

## Supplemental material

Supplemental Table 1 (intended for publication)

## Funding support

Renata Medical sponsored this study.

## Abbreviations

CHD: congenital heart disease
CoA: coarctation of the aorta
LPA/RPA: left/right pulmonary artery
PAS: pulmonary artery stenosis

---

**Supplemental Table 1:**
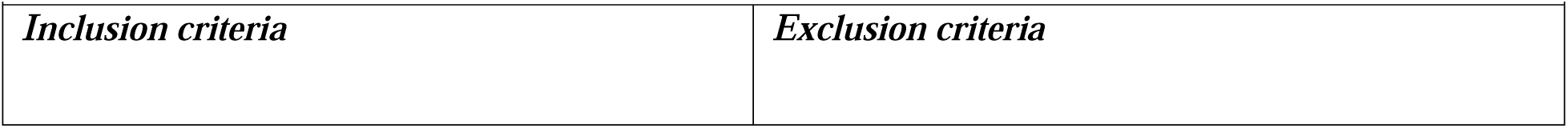
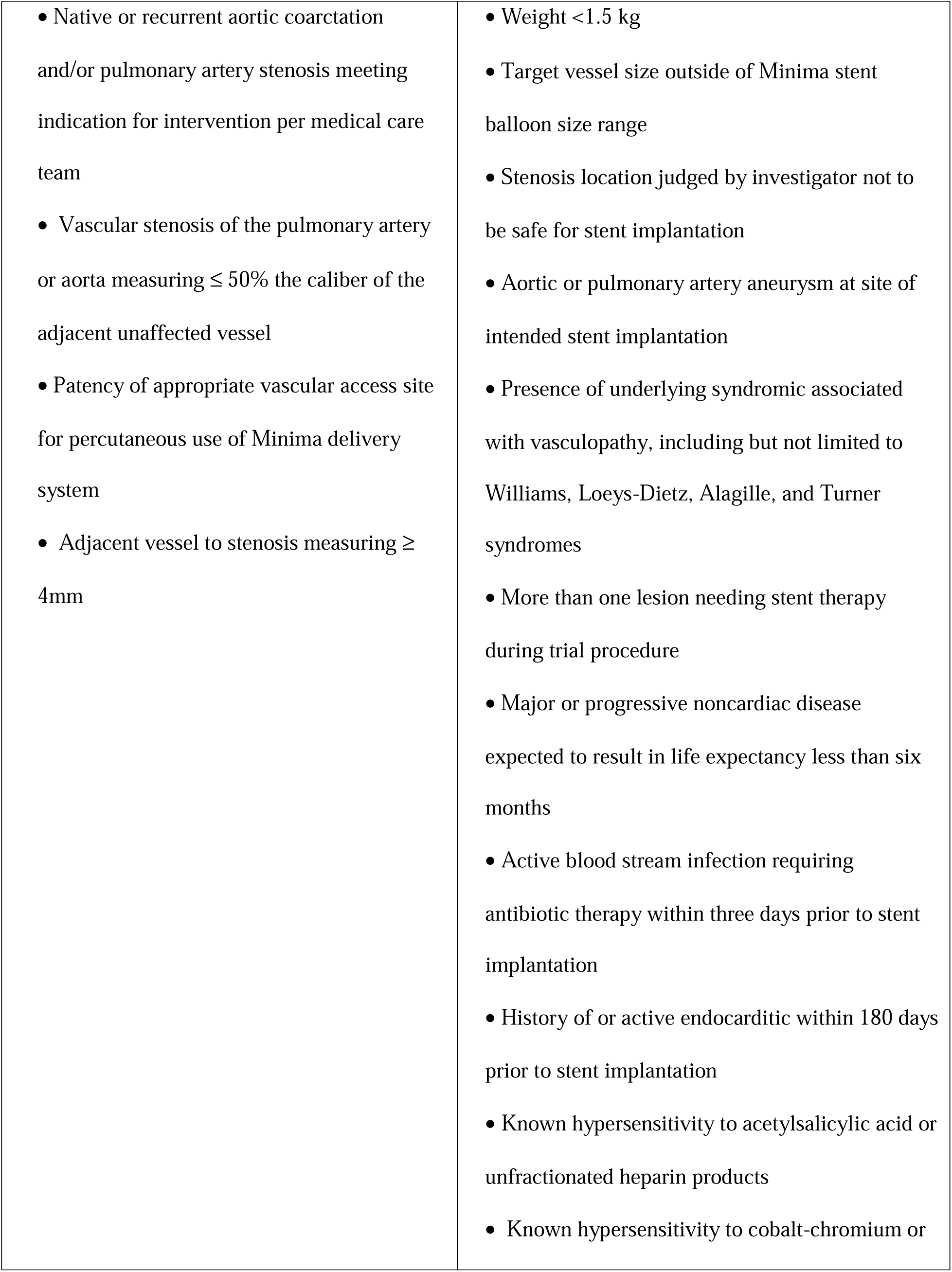
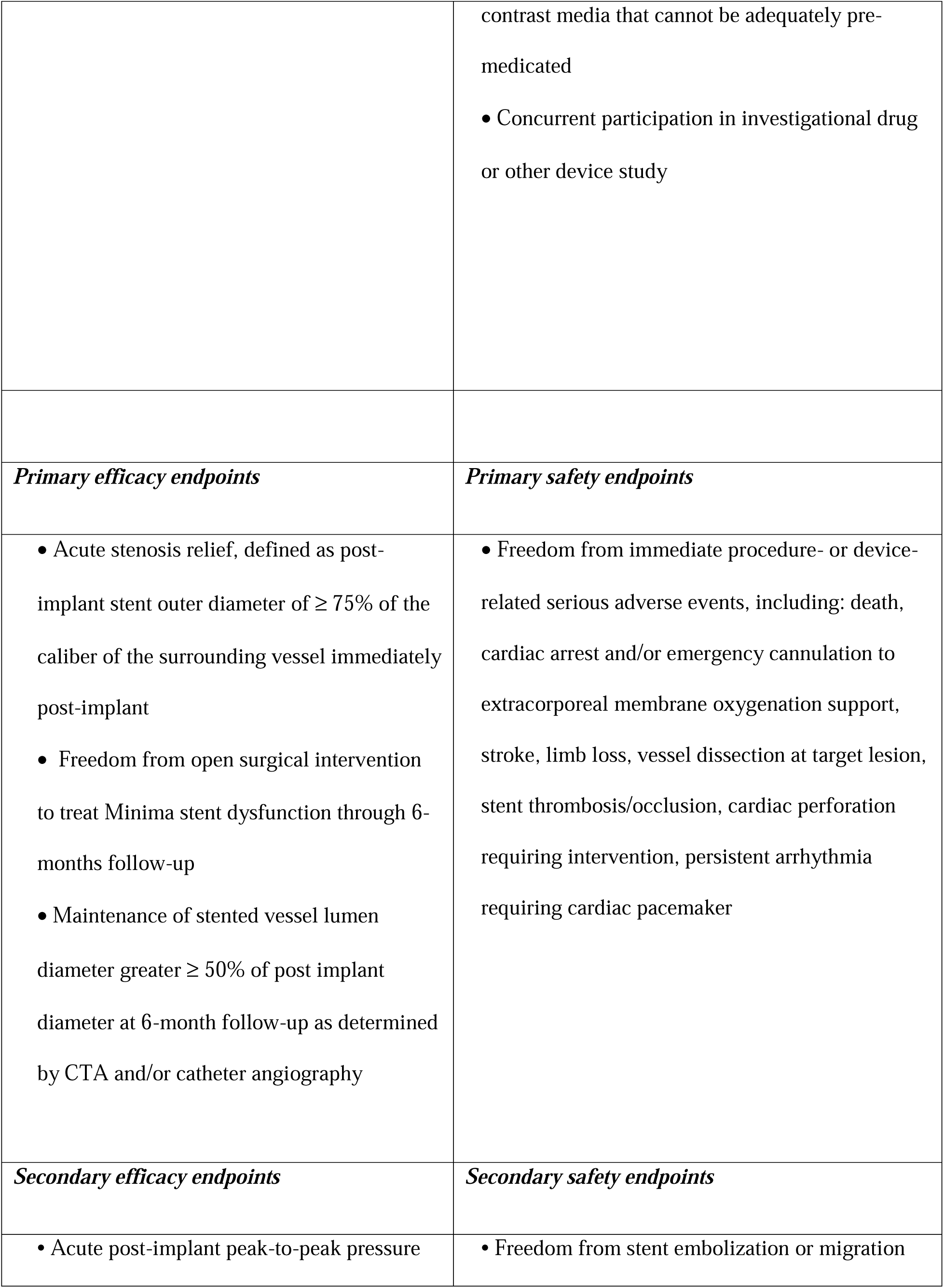
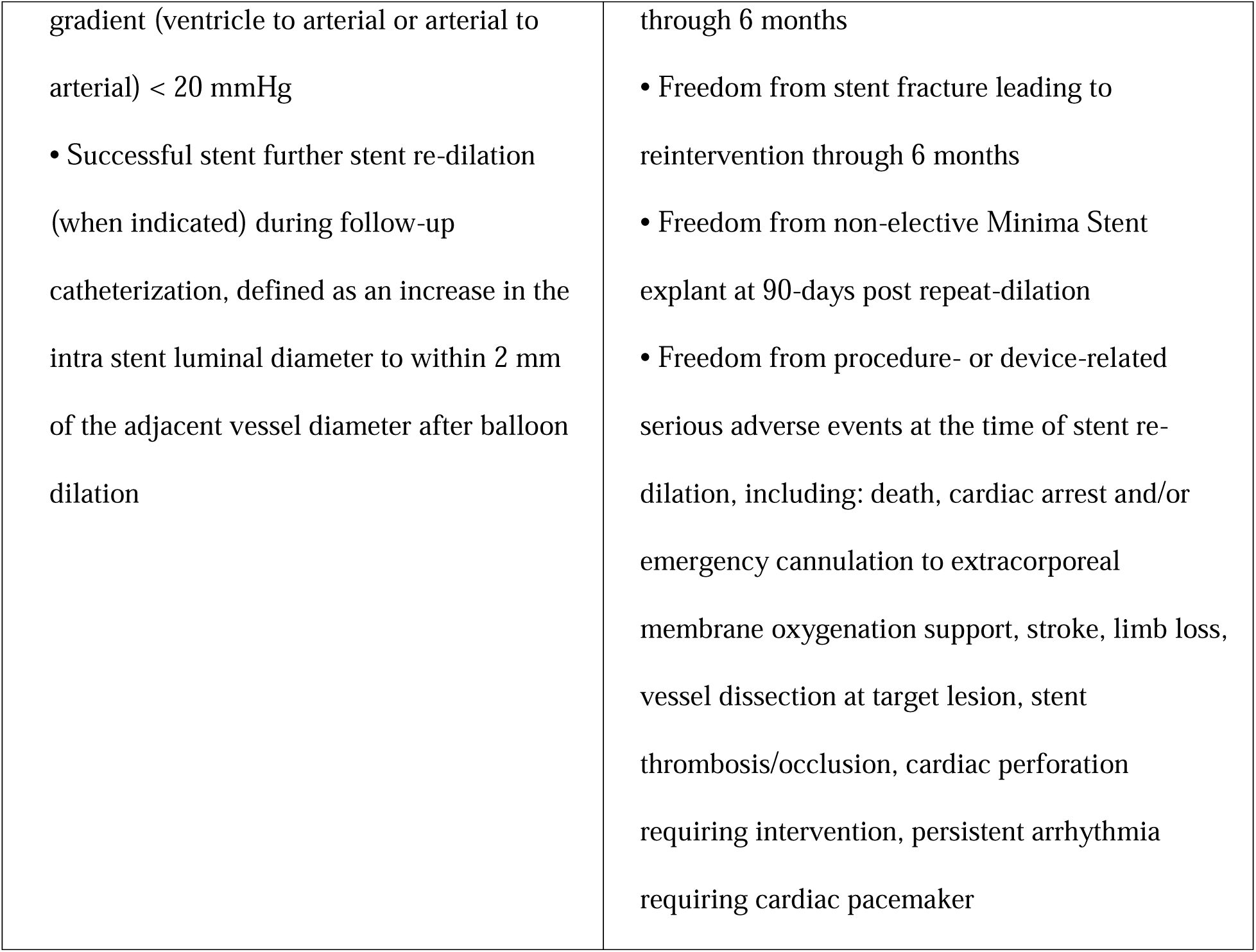
Inclusion and exclusion criteria for trial participation and endpoints for device efficacy and safety

## Notes

### Clinical Trial

NCT05086016

### Funding Statement

This study was funded by Renata Medical.

### Author Declarations

The IRB of Children's Hospital Los Angeles gave ethical approval for this work. The IRB of Cedars-Sinai Medical Center, Los Angeles, gave ethical approval for this work. The IRB of the University of Tennessee Health Science Center, in Memphis, TN, gave ethical approval for this work. The IRB of Seattle Children's Hospital gave ethical approval for this work. The IRB of Cincinnati Children's Hospital gave ethical approval for this work. The IRB of Nationwide Children's Hospital gave ethical approval for this work. The IRB of Children's Hospital Boston gave ethical approval for this work.

## References

1. O’Laughlin MP, Slack MC, Grifka RG, Perry SB, Lock JE, Mullins CE. Implantation and intermediate-term follow-up of stents in congenital heart disease. Circulation. 1993;88:605–14.

2. Okubo M, Benson LN. Intravascular and intracardiac stents used in congenital heart disease. Curr Opin Cardiol. 2001;16:84–91.

3. Ing FF, Khan A, Kobayashi D, Hagler DJ, Forbes TJ. Pulmonary artery stents in the recent era: Immediate and intermediate follow-up. Catheter Cardiovasc Interv. 2014;84(7):1123–30.

4. Zablah JE, Morgan GJ. Pulmonary artery stenting. Intervent Cardiol Clin. 2019;8(1):33–46.

5. Holzer RJ, Gauvreau K, McEnaney K, Watanabe H, Ringel R. Long-Term Outcomes of the Coarctation of the Aorta Stent Trials. Circ Cardiovasc Interv. 2021;14(6):e010308.

6. Forbes TJ, Kim DW, Du W, Turner DR, Holzer R, Amin Z, Hijazi Z, Ghasemi A, Rome JJ, Nykanen D, et al. Comparison of surgical, stent, and balloon angioplasty treatment of native coarctation of the aorta. J Am Coll Cardiol. 2011;58(25):2664–2674.

7. Beekman RH 3rd, Duncan BW, Hagler DJ, Jones TK, Kugler JD, Moore JW, Jenkins KJ. Pathways to approval of pediatric cardiac devices in the United States: challenges and solutions. Pediatrics. 2009;124(1):e155–62.

8. Wunnava S, Miller TA, Narang C, Nathan M, Bourgeois FT. US Food and Drug Administration Approval of High-risk Cardiovascular Devices for Use in Children and Adolescents, 1977-2021. JAMA. 2022;328(6):580–582.

9. Hatai Y, Nykanen DG, Williams WG, Freedom RM, Benson LN. Endovascular stents in children under 1 year of age: acute impact and late results. Heart. 1995;74(6):689–695.

10. Stanfill R, Nykanen DG, Osorio S, Whalen R, Burke RP, Zahn EM. Stent implantation is effective treatment of vascular stenosis in young infants with congenital heart disease: acute implantation and long-term follow-up results. Catheter Cardiovasc Interv. 2008;71(6):831–41.

11. Boe BA, Armstrong AK, Janse SA, Loccoh EC, Stockmaster K, Holzer RJ, Cheatham SL, Cheatham JP, Berman DP. Percutaneous Implantation of Adult Sized Stents for Coarctation of the Aorta in Children ≤20 kg: A 12-Year Experience. Circ Cardiovasc Interv. 2021;14(2):e009399.

12. Jenkins KJ, Beekman RH, Vitale MG, Hennrikus WL. Off-label use of medical devices in children. Pediatrics. 2017;139(1):e20163439.

13. Sutherell JS, Hirsch R, Beekman RH III. Pediatric interventional cardiology in the United States is dependent on the off-label use of medical devices. Congenit Heart Dis. 2010;5(1):2–7.

14. Berman DP, Morray B, Sullivan P, Shahanavaz S, Zahn EM. Results of the multicenter early feasibility study (EFS) of the Renata Minima stent as treatment for branch pulmonary artery stenosis and coarctation of aorta in infants. Catheter Cardiovasc Interv. 2024;104(1):61–70.

15. Zahn EM, Abbott E, Tailor N, Sathanandam S, Armer D. Preliminary testing and evaluation of the Renata minima stent, an infant stent capable of achieving adult dimensions. Catheter Cardiovasc Interv. 2021;98(1):117–127.

16. Mullins CE, O’Laughlin MP, Vick GW 3rd, Mayer DC, Myers TJ, Kearney DL, Schatz RA, Palmaz JC. Implantation of balloon-expandable intravascular grafts by catheterization in pulmonary arteries and systemic veins. Circulation. 1988;77(1):188–99.

17. Angtuaco MJ, Sachdeva R, Jaquiss RD, Morrow WR, Gossett JM, Fontenot E, Seib PM. Long-term outcomes of intraoperative pulmonary artery stent placement for congenital heart disease. Catheter Cardiovasc Interv. 2011;77(3): 395–9.

18. Hill J, Qureshi AM, Worley S, Prieto LR. Percutaneous recanalization of totally occluded pulmonary veins after pulmonary vein isolation-intermediate-term follow-up. Catheter Cardiovasc Interv. 2013;82(4): 585–91.

19. Barron DJ, Ramchandani B, Murala J, Stumper O, De Giovanni JV, Jones TJ, Stickley J, Brawn WJ. Surgery following primary right ventricular outflow tract stenting for Fallot’s tetralogy and variants: rehabilitation of small pulmonary arteries. Eur J Cardiothorac Surg. 2013:44(4):656–662.

20. Peng LF, McElhinney DB, Nugent AW, Powell AJ, Marshall AC, Bacha EA, Lock JE. Endovascular stenting of obstructed right ventricle-to-pulmonary artery conduits: a 15-year experience. Circulation. 2006;113(22):2598–605.

21. Morray BH, McElhinney DB, Marshall AC, Porras D. Intentional Fracture of Maximally Dilated Balloon-Expandable Pulmonary Artery Stents Using Ultra-High-Pressure Balloon Angioplasty: A Preliminary Analysis. Circ Cardiovasc Interv. 2016;9(4):e003281.

22. Ewert P, Riesenkampff E, Neuss M, Kretschmar O, Nagdyman N, Lange PE. Novel growth stent for the permanent treatment of vessel stenosis in growing children: an experimental study. Catheter Cardiovasc Interv. 2004;62(4):506–10.

23. Sullivan PM, Liou A, Takao C, Justino H, Petit CJ, Salazar JD, Ing FF. Tailoring stents to fit the anatomy of unique vascular stenoses in congenital heart disease. Catheter Cardiovasc Interv. 2017;90(6):963–971.

24. Peters B, Ewert P, Berger F. The role of stents in the treatment of congenital heart disease: Current status and future perspectives. Ann Pediatr Cardiol. 2009;2(1):3–23.

25. Lefort B, Lachaud M, El Arid JM, Neville P, Soulé N, Guérin P, Chantepie A. Immediate and midterm results of balloon angioplasty for recurrent aortic coarctation in children aged<1 year. Arch Cardiovasc Dis. 2018;111(3):172–179.

26. Glatz AC, Keashen R, Chang J, Balsama LA, Dori Y, Gillespie MJ, Giglia TM, Raffini L, Rome JJ. Outcomes using a clinical practice pathway for the management of pulse loss following pediatric cardiac catheterization. Catheter Cardiovasc Interv. 2015;85:111–117.

27. Alexander J, Yohannan T, Abutineh I, Agrawal V, Lloyd H, Zurakowski D, Waller BR 3rd, Sathanandam S. Ultrasound-guided femoral arterial access in pediatric cardiac catheterizations: a prospective evaluation of the prevalence, risk factors, and mechanism for acute loss of arterial pulse. Catheter Cardiovasc Interv. 2016;88:1098–1107.

28. Patel ND, Sullivan PM, Takao CM, Badran S, Ing FF. Stent treatment of ostial branch pulmonary artery stenosis: initial and medium-term outcomes and technical considerations to avoid and minimise stent malposition. Cardiol Young. 2020;30(2):256–262.

